# Oral Collagen for the treatment of Dermal Atrophy: A systematic review of human trials

**DOI:** 10.1101/2020.07.15.20154120

**Authors:** Rhydian Howell-Morris

## Abstract

**BACKGROUND:** Dermal atrophy (DA) or skin “thinning” can cause a substantial impact on quality of life and, due to barrier function damage, further health problems including cutaneous infection, skin tears and lacerations from minor trauma, impaired wound healing and chronic dermal inflammation. Some dietary products are targeted at therapeutic and functional treatments for skin ageing (of which mild DA is a component); however, while dietary collagen is amongst the most popular, particularly in the form of collagen peptides (CPs), in contrast to reviews for both over-the-counter and under prescription topical treatments for DA (e.g. Tretinoin), there is no reviewed literature of human trials testing the efficacy of orally administered collagen treatments applicable to DA; hence this review.

**OBJECTIVE:** To review the literature and assess available randomised-controlled trials (RCTs) testing the efficacy of orally administered collagen treatments for any skin properties that relate to the pathophysiology of DA and suggest their potential for medical and general public use in treating DA.

**EVIDENCE REVIEW METHOD:** A PubMed search was conducted using “(collagen) AND (supplementation OR treatment) AND (skin OR dermis)”, after which titles and abstracts were screened to decide if they matched the inclusion criteria for review. Results were collected up to 1^st^ of August 2019 and no lower limit on the year of publication was set.

**MAIN RESULTS:** Five studies with a total of 430 participants were included for review, with participants aged 24-70. Four out of the five studies used female only participants. The five studies used orally administered CPs, with dosage ranging from 570 mg/d to 10 g/d, running from 8 weeks to 6 months and assessed a range of skin properties relevant to DA including dermal thickness, epidermal thickness, dermal density, dermal collagen content, dermal collagen density and dermal elasticity. One of the studies combined CPs with several antioxidant ingredients to form the treatment and the remaining 4 studies used CPs as the only active ingredient. Methods to control for potential confounders were implemented in most studies including limiting exposure to sun, implementing a pre-treatment period of 1 week or more that controlled the use of cosmetics and intake of certain medications, micronutrient supplements and nutraceuticals with those restrictions continuing for the duration of the study. Given the heterogeneity of outcome measures across studies, quantitative analysis of results was not possible. In summary, the study with the antioxidant combined supplement showed a significant improvement in dermal thickness; two of the studies showed improvement in dermal collagen or pro-collagen content; three of the studies showed improvement in dermal elasticity; three studies showed improvement in dermal density or dermal collagen density; and lastly, no human study was found with the stated objective of assessing CPs effect specifically on DA.

**CONCLUSION:** Although definitive mechanistic cause-effect conclusions could not be drawn from the existing studies, they are supportive of beneficial effects of oral CP intake for treating characteristics of dermal atrophy. Further elucidation of the exact mode(s) of action that the CP intake has on improving dermal thickness, dermal density, and other skin biomarkers is necessary, with larger studies including more finely divided experimental and dose-response groups.

In conclusion, rigorousness of the trials must be improved to establish a cause-effect relationship between the CP intake and the beneficial effects for the skin atrophy, however potential has been demonstrated.

## INTRODUCTION

The skin is the largest organ in the body, it provides sensitivity to our environment and is the first line of defence against external insults such as exposure to chemicals, UV radiation and pathogens. It is also a vital effector for maintaining homeostasis of our internal environment including helping to maintain temperature and hydration levels.

The skin is made of three main layers: the epidermis, the dermis below, and below that the hypodermis or subcutaneous tissue. The structure, strength and flexibility of the dermis is primarily maintained by collagen and elastin proteins, with fibrous collagen constituting the majority of the dermis and providing the mechanical strength.

Dermal atrophy (DA), or skin “thinning” is increasingly recognised as not only a cosmetic concern but a possible chronic health condition, as it can involve substantial decrease in quality of life and, due to barrier function damage, further health problems including cutaneous infection, skin tears and lacerations from minor trauma, impaired wound healing and chronic inflammation [1]. It is noted clinically by a decrease in the thickness of the skin, visible vasculature and easy bruising and it may be detected earlier by measuring epidermal thickness and overall skin thickness (dermal and epidermal) values.

The pathophysiology of DA is characterised by decrease in the production of dermal extracellular matrix (ECM) constituents, notably fibroblasts, collagen [2], hyaluronic acid (HA) [3], and/or a decrease in keratinocyte proliferation resulting in reduced barrier function [1][4]. In fact, any pathologies of the dermal fibroblast will significantly disrupt the balance of the ECM and its other constituents because it’s the fibroblasts that synthesise dermal collagen, elastic fibres and orchestrate the matrix structure overall [5].

There are two broad aetiologies for the pathophysiological changes observed in DA. One aetiology is intrinsic, due to senescence, where it is a component of the recently named Dermatoporosis [1], which involves a decrease in collagen production [2] and an increased expression of metalloproteinases (MMPs) that can degrade collagen [6], as well as further inhibit the synthesis of ECM components in the dermis [7]. Exposure to UV irradiation from the sun can play an accelerating role in skin ageing too (photoageing) [7]. The other aetiology is from long-term use of potent topical and systemic corticosteroids, which can inhibit type I, type III and IV collagen synthesis, inhibit dermal HA synthesis and increase the production of matrix metalloproteinases (MMPs) [8][4][9].

An increasing range of dietary products are marketed to improve skin health and firmness in general [10], and some are even targeted at therapeutic and functional treatments for skin ageing, including the mechanisms of dermal thinning involved, without explicitly advertising as targeting DA. Given that the ECM is abundantly composed of collagen fibres [11], and the skin-conscious public are increasing becoming aware of this, dietary forms of collagen are amongst the most popular functional foods for skin health and anti-ageing, in the form of collagen peptides (CPs). There is a history of human studies and reviews for topical treatments for skin conditions, including DA as a component of skin ageing, both for over-the-counter cosmetics and for under prescription topical treatments such as Tretinoin [12] and topical lactic acid [13]; however there is no review of the literature applied to dermal atrophy examining the human trials that investigate the efficacy of orally administered collagen as a treatment.

Collagen is a triple helix protein composed of three polypeptide alpha chains [14], which when heated denatures and becomes gelatin. Each alpha polypeptide is composed of repeating triplets of amino acids (Gly-X-Y)_n_, where X and Y are most likely to be hydroxyproline (Hyp) or proline (Pro) [14][15]. Collagen oligopeptides (COPs) are simplified, smaller peptides of just two or three collagen amino acids in length that are produced by enzymes from the hydrolysis of gelatin. Collagen oligopeptides (COPs), also called collagen peptides (CPs), are water soluble, low-molecular weight (< 5KDa), and can be partially absorbed into the blood through digestion [16][17], making them more bioavailable than whole collagen proteins. Figure 1 shows the serum levels of Hyp before and after ingestion of CPs. Certain COPs such as Pro-Hyp and Hyp-Gly have been shown to have bioactivity in skin fibroblasts [18][19]. Figure 2 shows the effect of Pro-Hyp oligopeptide on the growth of dermal fibroblasts. In addition, certain COPs have more generally been proven to stimulate dermal metabolism [6] and have a positive effect on skin properties [20]. Collagen in the form of CPs/COPs is also called collagen hydrolysate (CH), so herein these terms may be used interchangeably.

**Figure 1.**
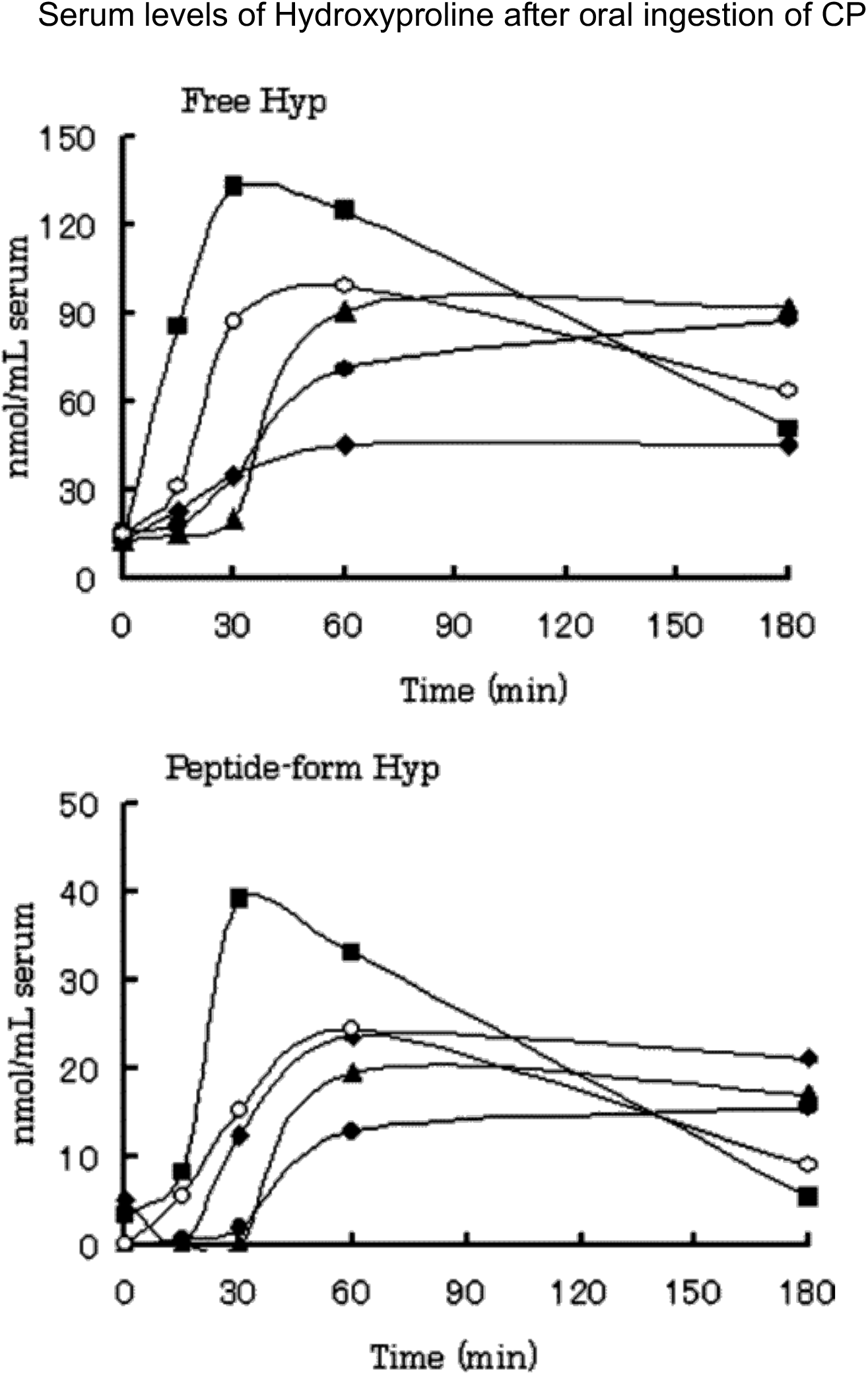
Serum levels of hydroxyproline in free (top) and peptide forms (bottom) after oral ingestion of the porcine type I gelatin hydrolysate (9.4 g). Data from all subjects are shown. Subjects, ○, 45 kg female 21 years old; ·, 45 kg female 23 years; ▪, 55 kg female 23 years; ▴, 80 kg male 39 years; ♦, 65 kg male 25 years. Reprinted (adapted) with permission from J. Agric. Food Chem. 2005, 53, 16, 6531-6536. Copyright 2005. American Chemical Society.

**Figure 2.**
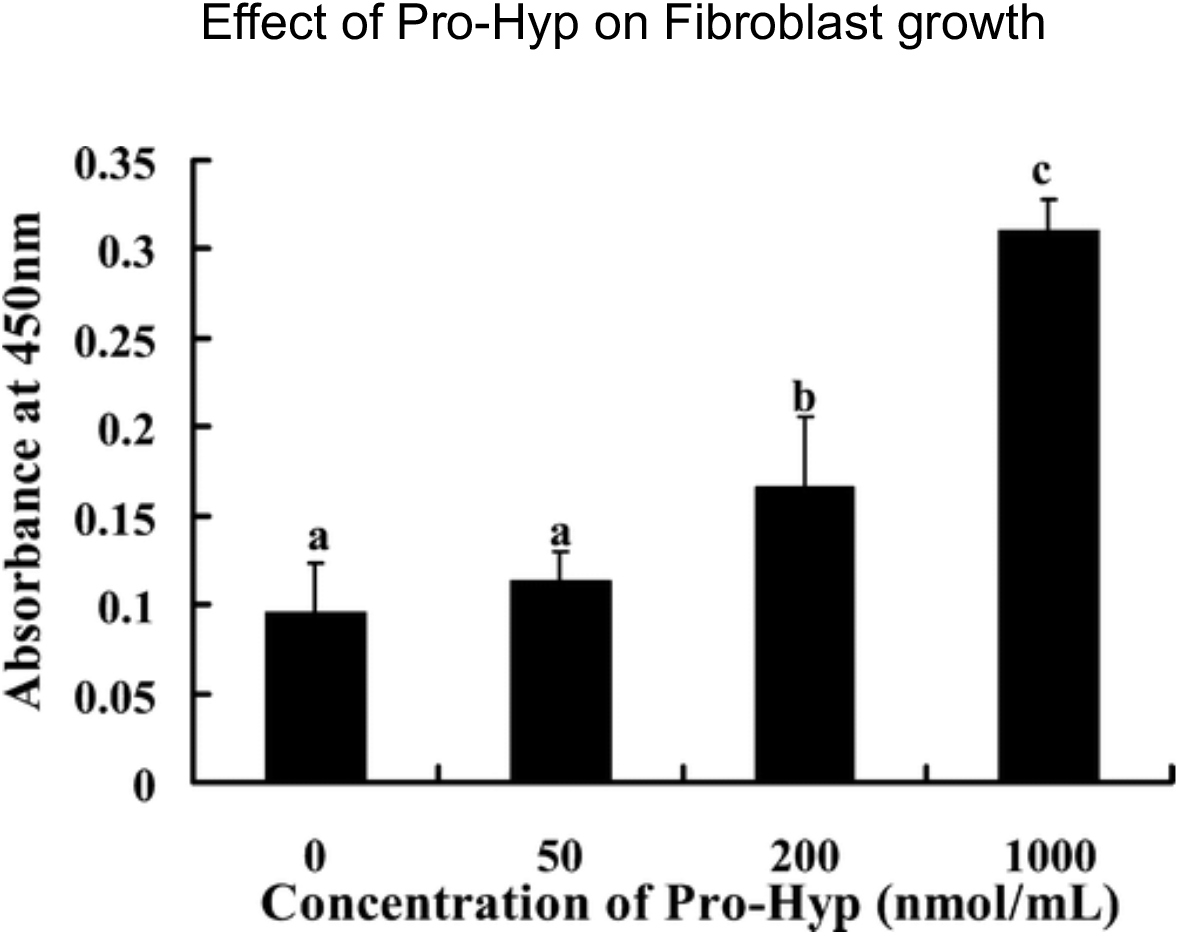
Dose response effect of Pro-Hyp on the growth of fibroblast on collagen gel in the medium containing 10% FBS. The mouse skin primary cultures of fibroblasts (5 × 103 cells) were cultured on a 96-well plate paved with collagen gel for 144 h. The data are shown as the mean ± SE; n = 8. The different letters on the values indicate significant difference (p < 0.05). Reprinted (adapted) with permission from J. Agric. Food Chem. 2009, 57, 2, 444-449. Copyright 2007. American Chemical Society.

There are a number of in vitro and animal studies that have shown the benefit that oral intake of CP has on skin health in general [21][22], its mode(s) of action [23], dose-dependent effects [24] and, even shown its direct effect on DA in a placebo-controlled study that demonstrated oral administration of CPs could correct skin atrophy in mice that were deficient in skin levels of the antioxidant superoxide dismutase (SOD) [25].

Superoxide dismutase deficiency causes DA by dysregulation of the extracellular matrix, through down-regulating mRNA for type I collagen gene (Col1a1), down-regulating mRNA for hyaluronan synthase 2 gene (Has2) and up-regulating the gene for metalloproteinase 2 (MMP2) [25]. In this study of SOD deficient mice, eight weeks of oral CPs treatment at 2% w/w dissolved in ultrapure water and orally administered via gavage (0.2 g/kg/day) was tested with transdermal co-treatment of 200 μL/day of a derivative of vitamin C at 0.1% w/w solution - having previously tested the effect of transdermal vitamin C alone [26]. The tested treatment was found to correct levels of mRNA in Col1a1 and Has2, but not MMP2, overall to partially normalise production of these ECM components and increase skin thickness.

In this review human studies of the effects CP supplementation have on human skin properties were summarised in the context of DA, for the first time to the best of my knowledge, and the potential they suggest oral CP intake has for treating DA in humans is evaluated.

### EVIDENCE REVIEW METHOD

A PubMed search was conducted using “(collagen) AND (supplementation OR treatment) AND (skin OR dermis)”, after which titles and abstracts were screened to decide if they matched the inclusion criteria. Once the screening was completed and the set of included studies was established, their full text was reviewed. The bibliographies of studies were also reviewed for further results. Results were collected up to 1st of August 2019 and no lower limit on the year of publication was set.

### INCLUSION CRITERIA

The included studies were randomised, controlled trials (RCTs) of any orally ingested collagen treatments in which the trial assessed the effects on DA or that assessed changes in particular skin properties that are pathophysiological in DA (such as changes in extracellular matrix components or changes in keratinocyte proliferation). Only studies written in English were included.

## RESULTS

### Study 1

In a randomised, double-blind, placebo-controlled clinical trial, 114 women (of which 108 completed the trial) aged 45-65 were randomly allocated to an experimental group (n = 57) and placebo group (n = 57), given 2.5 g of Bioactive CP (BCP) derived from porcine skin collagen type I in powder form, to be taken dissolved in liquid once daily for 8 weeks or given a placebo of maltodextrin [27]. Out of these participants, 48 were included for assessments (40 completed) by suction blister biopsies for quantitative analysis of levels of type I procollagen, elastin and fibrillin in skin fluid. Assessments suction blister biopsies were taken and made at baseline (week 0) and after 8 weeks of BCP intake. Samples were taken from the inner aspect of the right forearm.

Results of the suction blister biopsies for procollagen type I content showed an increase by 65% after 8 weeks of BCP treatment compared to the placebo group and was statistically significant (p < 0.01).

In addition to procollagen, BCP intake also promoted a statistically significant increase in elastin content by 18% in comparison to the placebo treatment after 8 weeks (p < 0.01).

BCP intake also led to an increase in fibrillin content of 6% in comparison to placebo after 8 weeks of treatment but this result failed to reach the level of statistical significance.

The authors concluded that oral intake of the BCP supplement (called Verisol®) for 8 weeks had positive impact on dermal ECM synthesis.

### Study 2

In another study involving Verisol®, a randomised, double-blind, placebo-controlled clinical trial of 105 women affected by cellulite (of which 97 completed the trial), aged 24-50, were randomly allocated to an experimental group (n = 53) and a placebo group (n = 52), given 2.5 g of Bioactive CP (BCP) derived from porcine skin collagen type I in powder (Verisol®), to be taken dissolved in liquid once daily for 6 months or given a placebo of maltodextrin to be taken by the same method by the placebo group [28].

Assessments were made at baseline, after 3 months of treatment and after 6 months of treatment, when measurements were taken of their dermal density by ultrasound and other skin properties not relevant to DA were assessed.

Results from assessments observed that dermal density of the placebo group fell over the 6 months from 6.85 to 6.45 (p < 0.01), dermal density of the experimental group rose over the 6 months from 7.29 to 7.66 (p <0.05), and relative dermal density between the groups was significantly improved in the experimental group compared with placebo (p < 0.05).

### Study 3

In another randomised, double-blind, placebo-controlled clinical trial, 106 women (of which 101 completed the trial), aged 40-65, were randomly allocated into an experimental group (n = 54) and placebo group (n = 52) and given 10 g of oral CPs of fish origin vs given a 10 g placebo, respectively, for 12 weeks [29].

Assessment was made of dermal collagen density, measured by high-frequency ultrasound at baseline, after 4 weeks and 12 weeks, and results showed an increase by 8.83% in the experimental group at 12 weeks vs no change in the control group (p = 0.007).

Alongside the clinical trial, an ex vivo experiment with the same participants involved an analysis of their skin explants incubated with three concentrations of Peptan. Measurements were taking for levels of moisture-retaining glycosaminoglycan (GAG) in the basal epidermis and for collagen content of the papillary dermis [29].

Results for levels of (GAG) showed a dose-dependent increase, although no change in transepidermal water loss (TEWL) was observed. Specifically at dosages of 0.01 mg/mL, 0.1mg/mL and 1 mg/mL, GAG increased in by 5x, 18x and 17x respectively, vs control (p < 0.001) with the authors concluding that saturation was achieved at some concentration between 0.1 mg/mL to 1 mg/mL to explain no further increase of GAG for the 1 mg/mL dosage.

Results for collagen content of the papillary dermis showed: at 0.01 mg/mL, a 3% increase; at 0.1 mg/mL, a 9% increase (p < 0.001); and at 1 mg/mL, a 5.4% increase, vs control (P < 0.01).

### Study 4

In a randomised, double-blind, placebo-controlled trial, 64 women (of which 53 completed the trial), aged 40-60, and diagnosed with photodamage wrinkled skin, were randomly allocated into an experimental group (n = 33) and a placebo group (n = 31), given a 50 mL supplement formulation containing a vehicle and 1 g of low-molecular weight collagen peptide (LMWCP) or a placebo formulation containing the vehicle and water instead of LMWCP [30]. Efficacy assessments were made at baseline, after 6 weeks and 12 weeks for changes in skin hydration, wrinkling, elasticity and an additional assessment at 12 weeks and 2 days for safety.

Results of assessments in skin elasticity showed overall elasticity was significantly higher in the experimental group vs the placebo group at 12 weeks (p = 0.025), as were values of net elasticity in the experimental group vs the placebo group at 12 weeks (p = 0.027). In absolute terms, the experimental group showed no significant increase in overall elasticity at week 12 vs its baseline value, but did show an increase in net elasticity at 12 weeks vs its baseline value (p = 0.002), whereas the placebo group showed no increase in net nor overall elasticity for any period during the trial.

The authors concluded that 1 g of LMWCP daily for 12 weeks significantly improved skin elasticity (as well as wrinkling and hydration that were assessed).

### Study 5

In a single-blind, controlled clinical-laboratory study, 41 participants, aged 35-70, were given a food supplement of 570mg marine CPs (MCPs), derived from deep sea fish combined with antioxidants comprised of 10mg grape-skin extract, 10mg of coenzyme Q_10_ of plant origin, 10mg of luteolin and 0.05mg of selenium of plant origin, twice daily over the course of 2 months [31]. Assessments were made 2 months prior to treatment, at baseline and at the end of 2 months of treatment, allowing the study to re-use the same participants as a control group (pre-treatment) and then experimental group. Assessments were made of facial skin physiology properties, including skin elasticity, by instruments and skin structure properties by ultrasound including epidermal and dermal thickness plus epidermal and dermal ultrasonic density.

Results from ultrasonic dermal thickness showed an increase from similar 3884*μ*m (+/- 30) pre-treatment and 3900*μ*m (+/- 31) at baseline readings compared to 4133*μ*m (+/-28), after treatment. Although the mean increase was noted from a small number of participants (n = 11), the differences between experimental and control group were statistically significant (p < 0.05). Dermal density also showed a statistically significant increase from 5.2 +/- 0.2 (pre-treatment) and 5.1 +/- 0.2 (baseline) compared to 6.3 +/- 0.1 after treatment vs control (p < 0.05).

For ultrasonic epidermal thickness and density results, there were no statistically significant changes after treatment vs before.

The skin physiological results showed a statistically significant increase in skin elasticity after treatment, from 34.06 +/- 1.54 pre-treatment and 33.66 +/- 1.21 at baseline compared to 40.26 +/- 0.87 after treatment (p < 0.0001). Skin moisture levels, however showed no statistically significant change after treatment compared to before.

Results from all properties measured showed a general stability between the pre-treatment assessment and baseline assessment, with no significant changes during that 2 month control period, allowing for conclusions to be drawn from changes at the end of treatment vs before as being caused by the intervention of the supplement taken.

## DISCUSSION

The incidence of dermal atrophy is growing, since it is a physiological component of skin ageing, combined with an ever-increasing ageing population, as well as it is having pathological extremes in dermatoporosis [32], photoaging[1] and from long-term use of potent corticosteroids [8][4]. While management of DA is becoming a growing challenge, there is also an increasing demand for natural and nutritional based therapies with low adverse effects [10].

One such potential functional food is collagen peptide (CP), or collagen hydrolysate (CH), that has been shown to be safe in multiple investigations [33][20]. Despite that safety and the fact that collagen makes up at least 70% of the dry weight of ECM [30][11], the efficacy of CP as a treatment for DA has yet to be reviewed. Therefore, the existing human studies of the effects CP has on the DA are the focus of this review.

This review tried to compile the most relevant human trials assessing oral CPs effects on DA and human trials of CPs related to skin properties that are pathophysiological in DA, yet only the latter could be found published up to 1^st^ of August 2019. As a result, the available studies were reviewed for their applicability to the therapeutic mechanisms of DA. It’s known that therapeutic aims of treating DA include improving dermal thickness, dermal density [25] and epidermal barrier function including through improved keratinocyte proliferation [3], so studies assessing outcomes corresponding to these mechanisms after intake of CP were those of significance to this review.

From those available studies matching the inclusion criteria, the context of their outcomes varied. Despite that for any study to be included skin properties involved in the DA were necessarily being assessed, because treatment for DA itself was not the central context of those studies, there were challenges involved from the heterogeneity of study designs encountered. All studies reviewed also necessarily used CPs as a supplement, whether as the only active ingredient or combined with others, yet many of the studies used different dosages from as little as 570 mg daily to as much as 10 g daily, making it difficult to compare studies and have cumulative or repeating evidence of dose-response effects on skin properties of DA.

The most important challenge for some of these human trials is the low number of participants, either in total or in particular inclusion sub-groups for certain assessments, leading to low statistical significance overall despite that there was usually high statistical significance in differences between experiment and control groups.

Only one study shed some light on its possible mode of action of CP intake with regard to improving dermal thickness [27], by assessing procollagen content as one of the biomarkers. As procollagen is a pre-cursor in the production of collagen [34], the resulting changes observed is surrogate for changes in dermal collagen synthesis, with results showing an increase after CP intake. Overall, the results of the biopsies observed in this study demonstrated a stimulatory impact of CP intake on the dermal ECM and the influence on collagen synthesis is a mechanism for attenuating specific dermal atrophy cases where the pathophysiology involves reduced dermal collagen synthesis.

In the study by Schunck et al. [28], the increased dermal density observed after treatment with the Bioactive CP supplement demonstrated that the dermis became more compact, which corresponds to strengthened connective tissue and potentially increased dermal collagen deposition.

In the study by Asserin et al. [29], the increased dermal collagen density observed after treatment with the supplement of CPs (called Peptan^®^F) and the ex vivo result showing an increase in collagen production in the papillary dermis suggest collagen synthesis might have been the mechanism through which the ingested CPs increased the dermal collagen density observed in the clinical trial, which is a therapeutic aim of DA treatment; however, the mode of action of Peptan was not actually demonstrated. The ex vivo experiments also showed an increase in GAG production, which could be significant since GAG production, such as hyaluronic acid production, can be reduced in dermal atrophy cases [35] resulting in reduced skin moisture [36], potential increased TEWL and decreased barrier function [3][36]. Also, given that hyaluronic acid is important for regulating dermal healing [37] and epidermal cell proliferation [38], an increase in GAG production from CP intake might be a therapeutic mechanism for DA cases that are characterised by decreased dermal hyaluronic acid production.

In the study by Kim et al. [30], the increased dermal elasticity observed after LMWCP intake offers another potential physiological mechanism for treating DA, as the production collagen and elastic fibre networks are correlated, both being synthesised by fibroblasts [5], and correlated in their degradation by MMPs. In addition, elastin protein is another important constituent of ECM for healthy, non-atrophied skin. However, the mode of action that LMWCP increased elasticity (whether an increase in production of elastin fibres, reduced inhibition of elastin synthesis or down- regulation of elastin-degrading MMPs) was not directly accounted for by any measurements in the trial.

In the study by De Luca et al. [31], the increased dermal thickness and density markers resulting from the CP supplement correspond to improved dermal collagen deposition and structure, which suggests CP intake has potential to attenuate these properties of DA, again depending on the unaccounted for mode of action and the particular therapeutic mechanism needed in the DA case. The results of this study showing an increase in dermal elasticity corresponds to either improvement in elastin production, reduced inhibition of elastin synthesis and/or reduced degradation of elastin by MMPs, so again depending on which mode(s) of action is responsible, there is potential for a normalising the corresponding mechanism involved in DA. However, compromising any significance and conclusions of cause-effect relationships regarding CPs in this study is the fact that the supplement was a multi-component formulation (called CELERGEN), therefore precluding inference as to what extent the marine CP specifically contributed to these improvements in skin thickness, isolated from the antioxidant ingredients. Further investigation would be required, separating various experimental sub-groups to compare the CELERGEN formulation to comparison formulations controlling for and evaluating each active ingredient.

In a final study that missed out on the inclusion criteria for not having a control group, 35 women (of which 31 completed the study) aged 35 to 65 with signs of skin ageing were given an oral supplement of CPs with vitamin C and two herbal extracts *H. sabdariffa*, and *A. chilensis*, of unknown quantities, to take with 200 mL of water once daily for 12 weeks [39]. Assessments were made of the dermal thickness by ultrasound at baseline, after 4 weeks, 8 weeks and 12 weeks.

Results of mean dermal thickness in the neck and cheekbone showed statistically significant increases. For skin in the neck area, after 4 weeks a mean increase of dermal thickness of 6.07% vs baseline was observed (p = 0.007), after 8 weeks a mean increase of 11.32% (p <0.001) and after 12 weeks of treatment a mean increase of 22.38% (p < 0.001). For skin in the cheekbone area, after 4 weeks an increase in dermal thickness of 5.81% vs baseline was observed (p = 0.001), after 8 weeks a mean increase of 14.69% (p <0.001) and after 12 weeks of treatment a mean increase of 24.97% was observed (p < 0.001).

Although this study had no control group and unspecified quantities for ingredients per administration, the size of the observed continual increases in dermal thickness give some statistical significance; however, given the mixture of active ingredients in the supplement, it is unknown to what extent the CP specifically caused these improvements in dermal thickness, especially as it is well established that vitamin C is also a co-factor in the synthesis of collagen [40] as well as an inhibitor of MMPs [41]. Nonetheless, the results of this study suggest that the supplement, whatever its constituents’ relative contributions, significantly improved dermal thickness, which is a therapeutic aim of treatments for DA.

## CONCLUSIONS

These studies have shown CH to be a potentially effective functional food applicable to the therapeutic aims involved in attenuating DA, however their broad applicability depends on their exact mode(s) of action, for which conclusive evidence is lacking in most cases. Further research is required to elucidate the cause-effect mechanisms at the molecular level between CP intake and the observed increases in dermal thickness, dermal density, collagen density, and dermal elasticity, as well as more research into the cause-effect relationship between different doses of the supplements.

Assessments that account for dose-response changes in molecular biomarkers such as gene expression for the synthesis of ECM constituents, for example, or dose-response changes in the gene expression of MMPs synthesis would provide the mechanistic evidence needed to explain the changes in macroscopic skin characteristics observed. Therefore, given the encouraging preliminary observations, human trials that include the aforementioned level of rigour are needed to improve the scientific basis that would verify oral CPs as worthwhile purchases by the general public for improving their DA and before their medical use could be recommended.

## Data Availability

All data is secondary from primary studies cited within the review manuscript.

## REFERENCES

1. Kaya G, Saurat JH. Dermatoporosis: a chronic cutaneous insufficiency/fragility syndrome. Clinicopathological features, mechanisms, prevention and potential treatments. Dermatology. 2007;215(4):284–94.

2. Varani J, Dame MK, Rittie L, Fligiel SE, Kang S, Fisher GJ, Voorhees JJ. Decreased collagen production in chronologically aged skin: roles of age-dependent alteration in fibroblast function and defective mechanical stimulation. Am J Pathol. 2006 Jun;168(6):1861–8.

3. Kaya G, Tran C, Sorg O, Hotz R, Grand D, Carraux P, Didierjean L, Stamenkovic I, Saurat JH. Hyaluronate fragments reverse skin atrophy by a CD44-dependent mechanism. PLoS Med. 2006 Dec;3(12):e493.

4. Schoepe S, Schäcke H, May E, Asadullah K. Glucocorticoid therapy-induced skin atrophy. Exp Dermatol. 2006 Jun;15(6):406–20.

5. Makpol S, Azura Jam F, Anum Mohd Yusof Y, Zurinah Wan Ngah W. Modulation of collagen synthesis and its gene expression in human skin fibroblasts by tocotrienol-rich fraction. Arch Med Sci. 2011 Oct;7(5):889–95.

6. Zague V, de Freitas V, da Costa Rosa M, de Castro GÁ, Jaeger RG, Machado-Santelli GM. Collagen hydrolysate intake increases skin collagen expression and suppresses matrix metalloproteinase 2 activity. J Med Food. 2011 Jun;14(6):618–24.

7. Fisher GJ, Kang S, Varani J, Bata-Csorgo Z, Wan Y, Datta S, Voorhees JJ. Mechanisms of photoaging and chronological skin aging. Arch Dermatol. 2002 Nov;138(11):1462–70

8. Measurement of dermal atrophy induced by topical steroids using a radiographic technique. https://www.ncbi.nlm.nih.gov/pubmed/322693 https://onlinelibrary.wiley.com/doi/abs/10.1111/j.1365-2133.1977.tb06142.x?sid=nlm%3Apubmed

9. Hengge UR, Ruzicka T, Schwartz RA, Cork MJ. Adverse effects of topical glucocorticosteroids. J Am Acad Dermatol. 2006 Jan;54(1):1–15.

10. Pérez-Sánchez A, Barrajón-Catalán E, Herranz-López M, Micol V. Nutraceuticals for Skin Care: A Comprehensive Review of Human Clinical Studies. Nutrients. 2018 Mar 24;10(4)

11. Hopkinson I. Molecular components of the extracellular matrix. J Wound Care. 1992 May 2;1(1):52–54.

12. Weiss JS, Ellis CN, Headington JT, Tincoff T, Hamilton TA, Voorhees JJ. Topical tretinoin improves photoaged skin. A double-blind vehicle-controlled study. JAMA. 1988 Jan 22- 29;259(4):527–32

13. Smith WP. Epidermal and dermal effects of topical lactic acid. J Am Acad Dermatol. 1996 Sep;35(3 Pt 1):388–91.

14. Shoulders MD, Raines RT. Collagen structure and stability. Annu Rev Biochem. 2009;78:929–58.

15. Phanat Kittiphattanabawon, Sitthipong Nalinanon, Soottawat Benjakul, and Hideki Kishimura, “Characteristics of Pepsin-Solubilised Collagen from the Skin of Splendid Squid (Loligo formosana),” Journal of Chemistry, vol. 2015, Article ID 482354, 8 pages, 2015

16. Iwai K, Hasegawa T, Taguchi Y, Morimatsu F, Sato K, Nakamura Y, Higashi A, Kido Y, Nakabo Y, Ohtsuki K. Identification of food-derived collagen peptides in human blood after oral ingestion of gelatin hydrolysates. J Agric Food Chem. 2005 Aug 10;53(16):6531–6

17. Ohara H, Matsumoto H, Ito K, Iwai K, Sato K. Comparison of quantity and structures of hydroxyproline-containing peptides in human blood after oral ingestion of gelatin hydrolysates from different sources. J Agric Food Chem. 2007 Feb 21;55(4):1532–5

18. Shigemura Y, Iwai K, Morimatsu F, Iwamoto T, Mori T, Oda C, Taira T, Park EY, Nakamura Y, Sato K. Effect of Prolyl-hydroxyproline (Pro-Hyp), a food-derived collagen peptide in human blood, on growth of fibroblasts from mouse skin. J Agric Food Chem. 2009 Jan 28;57(2):444–9

19. Shigemura Y, Akaba S, Kawashima E, Park EY, Nakamura Y, Sato K. Identification of a novel food-derived collagen peptide, hydroxyprolyl-glycine, in human peripheral blood by pre-column derivatisation with phenyl isothiocyanate. Food Chem. 2011 Dec 1;129(3):1019–24

20. Proksch E, Segger D, Degwert J, Schunck M, Zague V, Oesser S. Oral supplementation of specific collagen peptides has beneficial effects on human skin physiology: a double-blind, placebo-controlled study. Skin Pharmacol Physiol. 2014;27(1):47–55.

21. Matsuda N, Koyama Y, Hosaka Y, Ueda H, Watanabe T, Araya T, Irie S, Takehana K. Effects of ingestion of collagen peptide on collagen fibrils and glycosaminoglycans in the dermis. J Nutr Sci Vitaminol (Tokyo). 2006 Jun;52(3):211–5.

22. Tanaka M, Koyama Y, Nomura Y. Effects of collagen peptide ingestion on UV-B-induced skin damage. Biosci Biotechnol Biochem. 2009 Apr 23;73(4):930–2.

23. Zague V, de Freitas V, da Costa Rosa M, de Castro GÁ, Jaeger RG, Machado-Santelli GM. Collagen hydrolysate intake increases skin collagen expression and suppresses matrix metalloproteinase 2 activity. J Med Food. 2011 Jun;14(6):618–24.

24. Song H, Meng M, Cheng X, Li B, Wang C. The effect of collagen hydrolysates from silver carp (Hypophthalmichthys molitrix) skin on UV-induced photoaging in mice: molecular weight affects skin repair. Food Funct. 2017 Apr 19;8(4):1538–1546.

25. Shibuya S, Ozawa Y, Toda T, Watanabe K, Tometsuka C, Ogura T, Koyama Y, Shimizu T. Collagen peptide and vitamin C additively attenuate age-related skin atrophy in Sod1-deficient mice. Biosci Biotechnol Biochem. 2014;78(7):1212–20.

26. Murakami K, Inagaki J, Saito M, Ikeda Y, Tsuda C, Noda Y, Kawakami S, Shirasawa T, Shimizu T. Skin atrophy in cytoplasmic SOD-deficient mice and its complete recovery using a vitamin C derivative. Biochem Biophys Res Commun. 2009 May 1;382(2):457–61.

27. Proksch E, Schunck M, Zague V, Segger D, Degwert J, Oesser S. Oral intake of specific bioactive collagen peptides reduces skin wrinkles and increases dermal matrix synthesis. Skin Pharmacol Physiol. 2014;27(3):113–9.

28. Schunck M, Zague V, Oesser S, Proksch E. Dietary Supplementation with Specific Collagen Peptides Has a Body Mass Index-Dependent Beneficial Effect on Cellulite Morphology. J Med Food. 2015 Dec;18(12):1340–8.

29. Asserin J, Lati E, Shioya T, Prawitt J. The effect of oral collagen peptide supplementation on skin moisture and the dermal collagen network: evidence from an ex vivo model and randomized, placebo-controlled clinical trials. J Cosmet Dermatol. 2015 Dec;14(4):291–301

30. Kim DU, Chung HC, Choi J, Sakai Y, Lee BY. Oral Intake of Low-Molecular-Weight Collagen Peptide Improves Hydration, Elasticity, and Wrinkling in Human Skin: A Randomized, Double-Blind, Placebo-Controlled Study. Nutrients. 2018 Jun 26;10(7).

31. De Luca C, Mikhal’chik EV, Suprun MV, Papacharalambous M, Truhanov AI, Korkina LG. Skin Antiageing and Systemic Redox Effects of Supplementation with Marine Collagen Peptides and Plant-Derived Antioxidants: A Single-Blind Case-Control Clinical Study. Oxid Med Cell Longev. 2016;2016:4389410.

32. Dyer JM, Miller RA. Chronic Skin Fragility of Aging: Current Concepts in the Pathogenesis, Recognition, and Management of Dermatoporosis. J Clin Aesthet Dermatol. 2018 Jan;11(1):13–18. Epub 2018 Jan 1.

33. Liang J, Pei XR, Wang N, Zhang ZF, Wang JB, Li Y. Marine collagen peptides prepared from chum salmon (Oncorhynchus keta) skin extend the life span and inhibit spontaneous tumor incidence in Sprague-Dawley Rats. J Med Food. 2010 Aug;13(4):757–70.

34. Wu M, Crane JS. Biochemistry, Collagen Synthesis. 2019 Jan;

35. Kaya G, Rodriguez I, Jorcano JL, Vassalli P, Stamenkovic I. Selective suppression of CD44 in keratinocytes of mice bearing an antisense CD44 transgene driven by a tissue-specific promoter disrupts hyaluronate metabolism in the skin and impairs keratinocyte proliferation. Genes Dev. 1997 Apr 15;11(8):996–1007

36. Stern R, Maibach HI. Hyaluronan in skin: aspects of aging and its pharmacologic modulation. Clin Dermatol. 2008 Mar-Apr;26(2):106–22.

37. Monslow J, Sato N, Mack JA, Maytin EV. Wounding-induced synthesis of hyaluronic acid in organotypic epidermal cultures requires the release of heparin-binding egf and activation of the EGFR. J Invest Dermatol. 2009 Aug;129(8):2046–58.

38. Karvinen S, Pasonen-Seppänen S, Hyttinen JM, Pienimäki JP, Törrönen K, Jokela TA, Tammi MI, Tammi R. Keratinocyte growth factor stimulates migration and hyaluronan synthesis in the epidermis by activation of keratinocyte hyaluronan synthases 2 and 3. J Biol Chem. 2003 Dec 5;278(49):49495–504

39. Addor FAS, Cotta Vieira J, Abreu Melo CS. Improvement of dermal parameters in aged skin after oral use of a nutrient supplement. Clin Cosmet Investig Dermatol. 2018;11:195–201.

40. Pinnel SR, Murad S, Darr D. Induction of collagen synthesis by ascorbic acid. A possible mechanism. Arch Dermatol. 1987 Dec;123(12):1684–6. doi: 10.1001/archderm.123.12.1684

41. Farris PK. Topical vitamin C: a useful agent for treating photoaging and other dermatologic conditions. Dermatol Surg. 2005 Jul;31(7 Pt 2):814-7; discussion 818.

